# Ad-hoc Assembly of Lean Extracorporeal Membrane Oxygenation Systems for COVID-19

**DOI:** 10.1101/2020.04.12.20061929

**Authors:** Patrick Hunziker, Urs Zenklusen

## Abstract

**Background:** The COVID-19 epidemic is overwhelming intensive care units with bilateral pneumonia patients requiring respiratory assistance. Bottlenecks in availability of ventilators and extracorporeal membrane oxygenation may contribute to mortality, implying ethically difficult rationing decisions. It is unclear if accelerated equipment production will meet demand, calling for fallback solutions for life support in worst-case scenarios.

**Methods:** Veno-venous extracorporeal gas exchange (VV-ECMO) can provide vital support in bilateral lung failure. VV-ECMO essentially comprises large flow venous accesses, membrane gas exchange, and a blood pump. As thousands of FDA and CE certified Impella blood pumps and consoles are distributed globally for cardiac support, we explored ad-hoc assembly of lean ECMO systems by embedding Impella pumps coaxially in tubes in combination with standard gas exchangers.

**Results:** Ad-hoc integration of Impella blood pumps with gas exchange modules, standard cannulas for large bore venous access, regular ECMO tubing, Y-pieces and connectors led to lean ECMO systems with stable performance over several days. Oxygenation of 2.5-5 L of blood/minute is realistic. Benefit/risk analysis appears favorable if a patient requires respiratory support but cannot be supported because of lack of ventilators or unavailability of a required ECMO system.

**Conclusion:** Ad-hoc assembly of veno-venous ECMOs using Impella pumps is feasible and results in stable blood flow across gas exchange modules. However, such off-label use of the devices calls for specific ethical and regulatory considerations prior to their use as last resort in patients for whom no other treatment modalities are available.

## Background

In COVID-19,^1^ bilateral viral pneumonia^2^ is a leading cause of death.^3^ In intractable patients with severe hypoxemia viral pneumonia due to H1N1 influenza, a benefit of ECMO therapy for specific groups with severe disease was recorded ^4,5,6,7^, suggesting that ECMO support will play a role in COVID-19, too.

Global healthcare systems have not been well prepared for a pandemia with such a rapid spread of a highly infectious virus. The large numbers of infected individuals and the large number of patients requiring hospitalization, and in particular, intensive care for respiratory support is overwhelming regional health care systems. Key consumables like face masks, key infrastructures like intensive care beds and key systems for respiratory assist like ventilators are not everywhere available in sufficient numbers, leading to a buying frenzy and efforts for rapidly building large numbers of ICU ventilators. An insufficient number of resources may lead^8^, and already has led^9^, to rationing of potentially life-saving activities like ventilation. In some areas, large patient numbers and insufficient resources have already led to rationing of medical activities like Intensive Care Unit admission and ventilation by excluding some patients with high predicted mortality and reduced life expectation. This calls for rapid efforts for improving availability of vital support equipment^10^.

## Methods

The feasibility of ad-hoc integrating Impella blood pumps that are already on-site in many hospitals with standard ECMO gas exchange cartridges, tubes and connectors was tested using different connection techniques. Figure 1 shows the concept, the used parts, assembly process and the results of such lean ad-hoc ECMO systems. Prototype systems were built as a first simplified closed loop with an embedded Impella CP pump only, as a second closed loop setup incorporating a venous suction cannula, a venous return cannula, an oxygenator and in-line Impella pump driven by a standard Impella AIC console (Abiomed, Danvers MA, U.S.A), and a third system additionally containing a second gas exchange module flushed with nitrogen to simulate oxygen extraction in patients. Abiomed Impella CP and Impella 5.0 pumps, and a standard ECMO oxygenator (Hilite 7000, Stolberg, Germany) were used. Key steps are the coaxial introduction of the Impella pump into the tube through a Y tube connector, prevention of recirculation of the pumped blood around the Impella pump head by suited obstruction/constriction of the outer tube, suited tube diameter stepups/stepdowns to minimize flow resistance at pump inlet/outlet and the use of large enough venous drainage cannulas (17-29F; 24-31F dual lumen cannulas) and sufficiently large (17-22F) jugular venous return cannulas. Supplement table 1 is a list of components used and gives detailed tips and tricks for easy assembly.

**Table 1:** Benefit/risk analysis and minimization of risks.

| Potential patient benefit of “Ad-hoc ECMO” |  | Benefit improbable | Benefit possible |
| --- | --- | --- | --- |
|  |  | Good prognosis with available ventilatory support |  |
|  |  | Very high predicted mortality despite ECMO |  |
|  |  | Conventional ECMO devices available |  |
|  |  |  | Fallback option in ventilation failure: Need for ECMO with realistic survival probability, but ECMO unavailable |
|  |  |  | Primary therapy for lung failure: Need for ventilatory assist for survival, but ventilators unavailable |
| Potential Hazard | Probability | Risk | Risk minimization |
| Thrombosis, embolism | Low to moderate | Low, preventable | Maintain therapeutic anticoagulation<br>Visual control of tubes and gas exchange module. Change when necessary. |
| Air embolism | Low to moderate | Low, preventable | Avoid or securely close access port on low-pressure side of pump. Install pump on low-pressure side of gas exchanger. |
| Hemolysis | Moderate | Low | Drive pump at level necessary for reasonable oxygenation, not higher |
| Arterial complications | Low | Low, preventable | Vascular access by experienced physician, ultrasound guided vascular puncture. |
| Infection | Low-moderate | Low-moderate | Monitor inflammation markers, repeated blood cultures. Replace system/antibiotics. |

**Figure 1:**
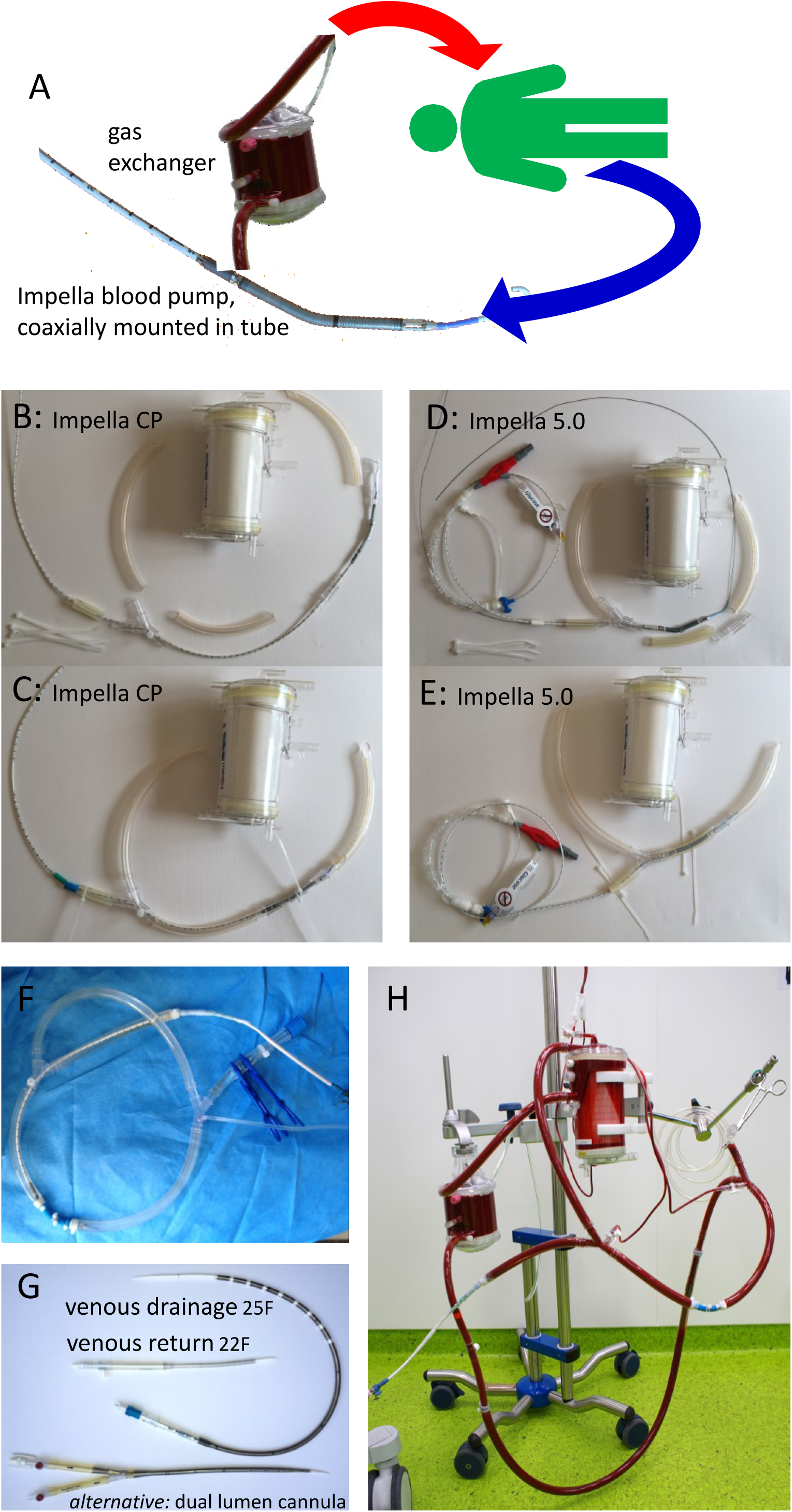
Ad-hoc ECMO. A: Basic connectivity of ad hoc ECMO composed from Impella pumps and standard oxygenators. B, C: pieces and assembly of Impella CP-based ECMO system. D,E: pieces and assembly of Impella 5.0-based ad-hoc ECMO system. F: closed-loop setup for pump testing, was run for 48 hours and showed stable performance. G: venous access cannulas; the 25F and 22F cannula were used in F. F: closed loop system including Impella pump, venous drainage and return cannulas, oxigenator module; to simulate oxygen extraction by a patient, an additional gas exchange module (flused with N2) is added. 48 hour – testing of this setup with reconstituted blood showed stable flows, and stable gas exchange.

Multiday test runs were done first in a simple closed loop system (Figure 1F) with saline and then in a complete system incorporating the Impella pump, venous drainage and return catheters and gas exchange modules with reconstituted blood from packed red blood cells and fresh frozen plasma at a hematocrit of 30% with heparin anticoagulation. Flows and pressures were determined using the sensors built into the Impella catheters/consoles and using external pressure sensors. Oxygenators were tested visually for clots formation and pressure gradients were determined over time. In the setup using two sequential gas exchange modules (the first flushed with N2 for partial deoxygenation) and the second with O2 for oxygenation, gas exchange over time was followed by blood gas analysis.

Figure 2a shows predicted, pressure-gradient dependent pump performance of the Impella pumps used, based on available Impella performance charts and shows observed system flow performance in our CP-based systems. Figure 2b shows expected pressure drops across typical cannulas according to publicly available data^11,12,13,14^ and gives the measured pressure drops across the used oxygenator type when driven at various flows using standard ECMO consoles in patients with a hematocrit of approximately 30%, based on own historical observations.

**Figure 2:**
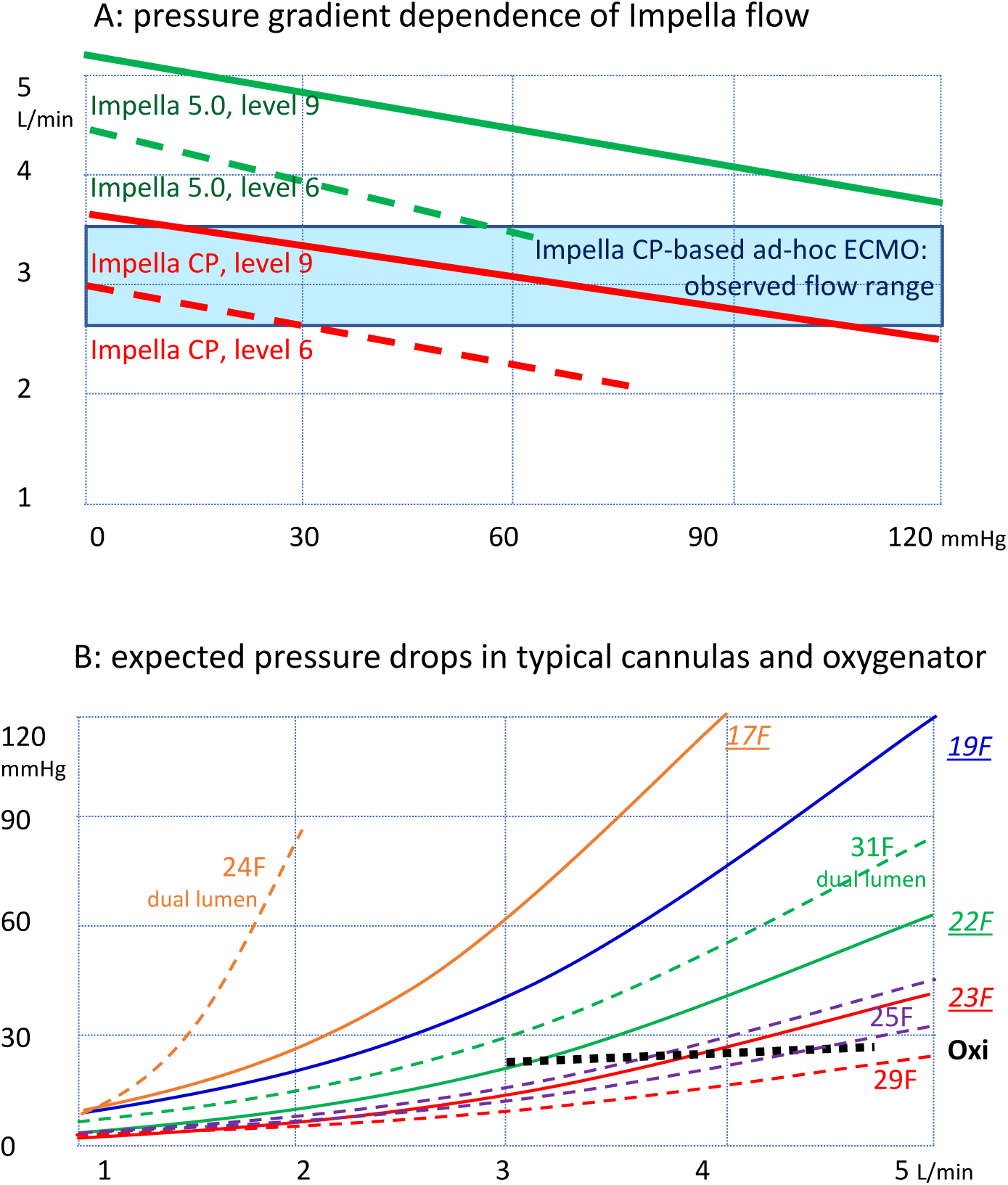
A: Flow-pressure relation prediction of used Impella CP and 5.0 pumps (red, green lines) based on performance chart. Observed flow range in assembled ad-hoc ECMO systems driven by Impella CP (blue). Observed flows were in the high range of the prediction, presumably benefiting from the improved flow characteristics of an axial pump positioned in a coaxial tube. B: Flow-pressure relation estimates of typical cannulas and oxygenator. Diverse data sources, diverse measurement conditions. Dotted black line/Oxi: observed pressure drops across gas exchange module Hilite 7000LT in patients with ECMO. Dashed lines: estimated pressure drops in venous drainage cannulas(55-65 cm length). Red: Maquet 29F; violet: Edwards QuickDraw 25F, Medtronic Biomedicus 25F; Green: Maquet Avalon 31 F bicaval (drainage lumen); orange: Medos Novaport dual lumen Twin24F (drainage lumen). Solid lines, underlined labels: estimated pressure drops in venous return cannulas (15-23cm length). Blue: Maquet 19F. green: Edwards 22F Red: Maquet/Getinge 23F cannula, yellow: 17F cannula.

Benefit/risk analysis was performed. This included patient risk and selection, probability of survival with/without added treatment, institutional setup and known risks of blood pumps including device thrombosis, embolism, infection, bleeding, hemolysis and air embolism.

## Results

Systems were successfully built using standard equipment with both, the Impella CP and the Impella 5.0 pump, although the different pump sizes require differently sized tubes and Y pieces. The Impella CP fit nicely through a standard 3/8” Y piece while the 5.0 Impella pump needs a larger Y piece and some special care to fit through the Y piece as described in more detail in the supplement.

Using the Impella CP, blood flows of up to 3.6L/min of saline (minimal circuit) and of 3.5L/min of blood (complete system) could be achieved at pump performance level 9: pressure gradients and flows across the system remained stable over 48 hours. Flows were at least as high as predicted by combining pump performance and component pressure drop charts. The combination of pump performance chart, cannula pressure drop chart and experimental observations indicates that when using large enough cannuals, the “sweet spot” for Impella CP based systems is between 2.8 and 3.5 L/min, while for Impella 5.0, it is between 3.5 and 4.5 L/min.

Given an oxygen binding capacity of 1.34ml/g hemoglobin, this translates to an oxygen transport capacity at a normal hemoglobin content of ECMO blood of up to 200ml/L × 3.5L/min, i.e. 700ml oxygen/min; at a hypothetical venous oxygen saturation of 60% with a near-normal body oxygen consumption of approximately 125ml/min/m^2^, even taking some venous/ECMO recirculation and some anemia into account, such an Impella CP-based ad-hoc ECMO will still result in a significant contribution to systemic oxygen delivery.

The gas exchange module showed no visual evidence of clots and the pressure gradient across the module remained unchanged over the observation time.

In the dual gas exchanger setup that uses a first gas exchange module in-line to partially deoxygenate the blood and then uses the second gas exchanger to re-oxygenate the blood, we observed a stable, near-maximal oxygen content after the second oxygenator over the observation time of 2 days, although the experiments performed at room temperatures and with N2 for O2 washout may not fully reflect physiological conditions.

Results of the Benefit/Risk analysis are shown in Table 1. We determined that the benefit/risk relation for using such an off-label system is most acceptable in a scenario with a patient who cannot be ventilated conventionally because a ventilator is not available or who requires an ECMO because of disease severity but standard ECMO systems are no more available.

## Discussion

Rapid assembly of an ad-hoc ECMO system is feasible using an Impella blood pump, e.g. the Impella CP or 5.0, a standard Abiomed Impella console, a standard ECMO gas exchange module and standard tubing/cannulas that are already widely distributed in hospitals, following the constructive approach and material delineated in this contribution. Assembling an ad-hoc ECMO system should pose no major problem for a trained perfusionist or a physician experienced in installing and using ECMO and Impella systems.

Such ad-hoc ECMO systems might prove to have a life-saving potential if used as a last-resort option in patients who urgently need gas exchange support due to bilateral COVID pneumonia where conventional ventilation fails but a conventional ECMO device is not rapidly available, and might offer hope as primary therapy in life-threating lung failure where ventilators are no more available. A patient benefit is improbable if the probability of outcome is anyway very good or very bad, or if standard ventilators and conventional ECMO devices are available that should be used preferentially. Experience in large-bore venous access and in handling blood pumps including ECMO and Impella is probably an important success factor for such an approach.

An advantage is the use of proven standard equipment like Impella consoles and Impella blood pumps, of which thousands are already distributed in many hospitals in the U.S. and Europe. In our institution, such Impella-based ad-hoc ECMO systems almost double the number of ECMO treatments that could be delivered to patients.

## Limitations

While certified for conventional hemodynamic support, Impella consoles and Impella pumps are not certified by regulatory bodies for such nonstandard use. While we saw that a trained perfusionist will easily master the technicalities of system composition, there will still be a learning curve in composition and handling such a system over time and questions remain regarding liability, formal training, etc. Before establishing and using such last-resort setups in individual patients, the physicians, perfusionists and institutions may thus benefit from consultation with ethical advisors and regulatory bodies. For a given patient, this may encompass a careful patient-centered assessment of potential benefit and risk, ethical and regulatory aspects, in particular examining the expanded access possibilities for medical devices in emergencies by FDA ^15^, the European Medical Agency ^16,17^ and others.^18^

The system has not yet been applied to patients in our hospital as the ventilation and ECMO infrastructure at our site at the time of writing is not yet exhausted and does therefore not call for application of last-resort measures, while the time window for a formal, pivotal clinical trials is too narrow.

In conclusion, composing ad-hoc ECMO systems by integrating readily available Impella blood pumps with standard oxygenators and off-the-shelf cannulas may offer another opportunity to oxygenate, to “recover the lungs” and hopefully to save lifes of patients with viral pneumonia when standard equipment is no longer available because demand exceeds availability.

## Data Availability

Data are given in supplement.

## Acknowledgments

Thanks to Christoph Nix, Abiomed, Danvers U.S.A. for helpful discussions and information about pressure/flow relations of Impella pumps.

## Supplement 1: key construction steps

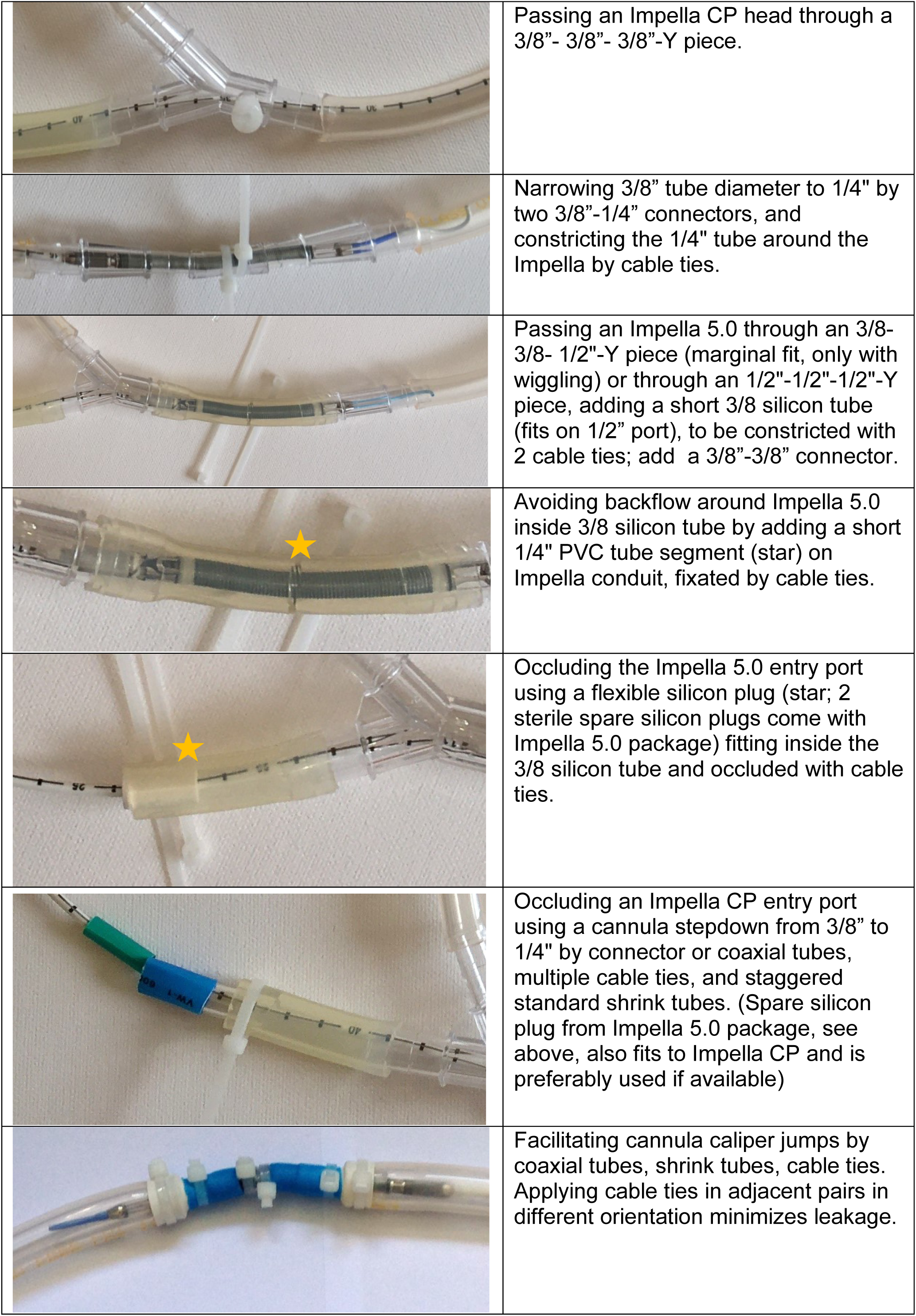

## Supplement 2: Materials, Compatibilities, Tricks &Tips

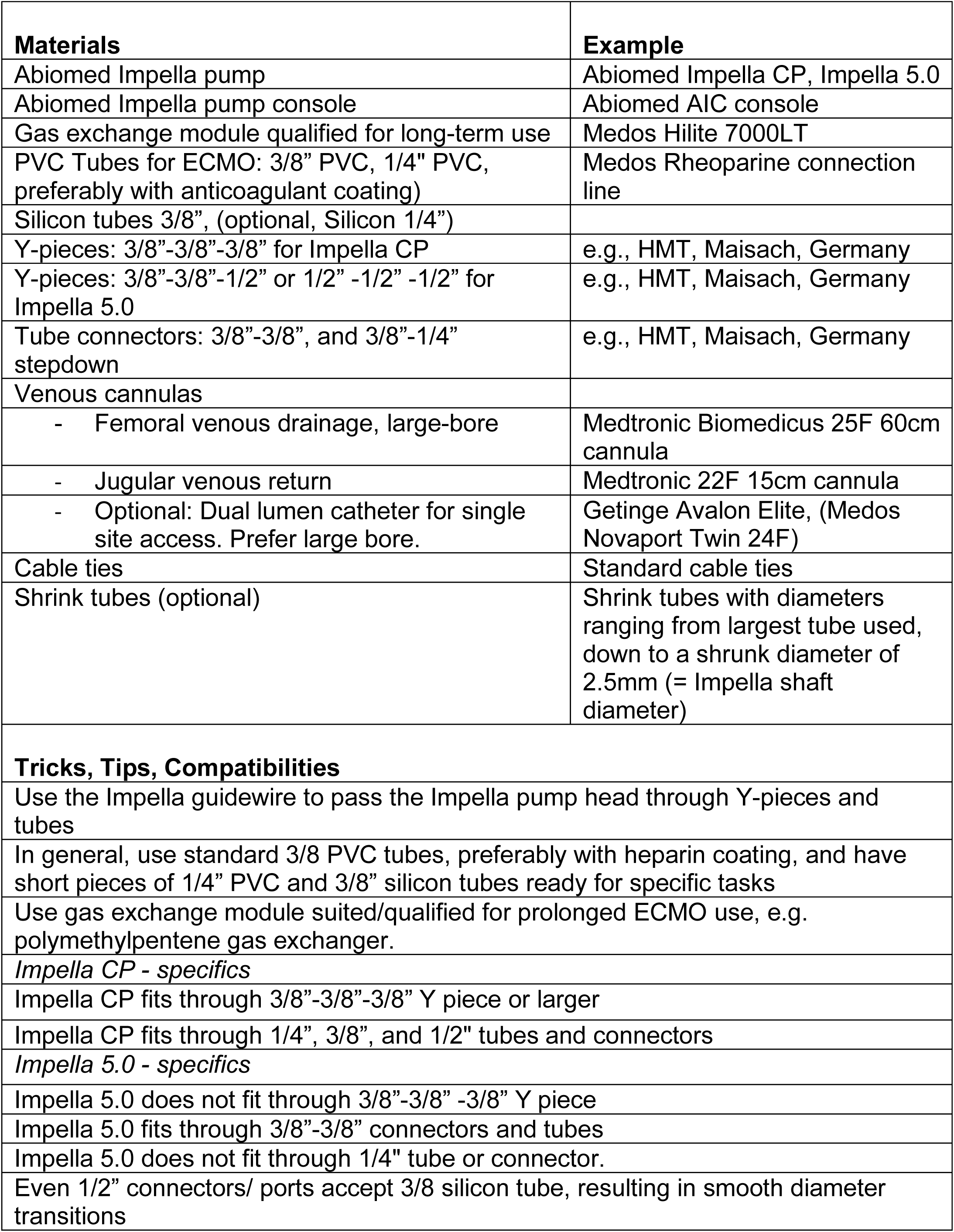

